# A Natural History of *NAA15*-related Neurodevelopmental Disorder Through Adolescence

**DOI:** 10.1101/2024.04.20.24306120

**Authors:** Rikhil Makwana, Carolina Christ, Rahi Patel, Elaine Marchi, Randie Harpell, Gholson J. Lyon

**Affiliations:** Department of Human Genetics, New York State Institute for Basic Research in Developmental Disabilities, Staten Island, New York, United States of America; George A. Jervis Clinic, New York State Institute for Basic Research in Developmental Disabilities, Staten Island, New York, United States of America; Biology PhD Program, The Graduate Center, The City University of New York, New York, United States of America

## Abstract

NAA15 is a member of the NatA N-terminal acetyltransferase complex, which also includes the NAA10 enzymatic sub-unit. Individuals with variants in the *NAA15* coding region develop *NAA15*-related neurodevelopmental syndrome, which presents with a wide array of manifestations that affect the heart, brain, musculoskeletal system, and behavioral and cognitive development. We tracked a cohort of 27 participants (9 females and 18 males) over time, each with a pathogenic *NAA15* variant, and administered the Vineland-3 assessment to assess their adaptive functioning. We found that the cohort performed significantly worse compared to the normalized Vineland values. On average, females performed better than males, and they performed significantly better on the Motor Domain and Fine Motor Sub-Domain portions of the assessment. Over time, females showed a decrease in adaptive functioning, with the decline being especially correlated at the Coping, Domestic, and Fine motor sub-domains. Males (after excluding one outlier) showed a moderate positive correlation between age and ABC standard score. Ultimately, additional longitudinal data should be collected to determine the validity of the between sex-differences and to better understand the change in adaptive behavioral outcomes of individuals with *NAA15*-neurodevelopmental disorder as they age.

## Introduction

N-terminal acetylation is a prevalent protein modification that involves the addition of an acetyl group to the alpha-amino group of the N-terminal amino acid of a protein (Aksnes et al., 2019). This enzymatic process is catalyzed by a group of N-terminal acetyltransferases (NATs) and plays a pivotal role in various cellular processes, including protein stability, protein-protein interactions, and cellular localization (Aksnes et al., 2015; Bienvenut et al., 2012; Dikiy & Eliezer, 2014). There are seven known NAT types, with NatA being the most prominent, believed to be responsible for modifying 40-50% of the human proteome (Deng et al., 2019; Feng & Ma, 2016; Van Damme, 2021). The NatA complex is composed of the catalytic sub-unit NAA10, and the auxiliary sub-units NAA15 and HYPK (Deng et al., 2019; Dörfel et al., 2017; Feng et al., 2016; Knorr et al., 2019; Weidenhausen et al., 2021).

Dysregulation of NAA10 and NAA15 expression or activity has been associated with the pathogenesis of various diseases, including cancer (Kuhns et al., 2018; Le et al., 2023; Zhang et al., 2020; Zhu et al., 2024), neurodevelopmental disorders (Cheng et al., 2018; Lyon et al., 2023; Makwana et al., 2024; Sidhu et al., 2017; Wu & Lyon, 2018), and cardiovascular diseases (Belbachir et al., 2023; Huth et al., 2023; Ritter et al., 2021; Støve et al., 2018; Ward et al., 2021). More notably, mutations in the X-linked gene *NAA10* lead to Ogden Syndrome, which has numerous effects on an individual’s development and overall health. *NAA15*-related neurodevelopmental syndrome has overlapping symptoms with Ogden Syndrome, including intellectual disability, atypical or delayed development, heart anomalies, and more features previously described, though patients with *NAA15* variants typically have milder phenotypes (Cheng et al., 2020; Dörfel & Lyon, 2015; Lyon et al., 2023; McTiernan et al., 2018; Myklebust et al., 2015; Patel et al., 2024; Pesz et al., 2018; Stessman et al., 2017; Støve et al., 2018; Umlai et al., 2021; Wong et al., 2019; Zhao et al., 2018). Additionally, *NAA15* variants are associated with musculoskeletal and neuromuscular abnormalities that can appear in childhood or may have a delayed appearance until adulthood (Danti et al., 2024; Monestier et al., 2019; Pesz et al., 2018; Tian et al., 2022; Yubero et al., 2022).

The aim of the current study is to further describe adaptive functioning in individuals with *NAA15*-related neurodevelopmental syndrome by analyzing prospective data to understand how this disorder progresses over time.

## Methods

### Participants

All families that participated in the investigation signed Institutional Review Board (IRB)-approved consents and HIPAA forms. The scope of the project was explained in lay language. The participant population included children with *NAA15* pathogenic variants that the principal investigator has previously worked with for other research projects. Participants were not compensated for their time. Overall, there were 27 families who were administered the assessment. Probands were assigned NAA15-XXX designations based on an internal registry known only to the researchers to ensure anonymity and confidentiality while allowing unique probands to be tracked in the literature.

### Cognitive Assessment

The parents of the participants were administered Vineland-3 which is composed of three core domains: Communication Daily Living Skills, and Socialization. These domains are further split into sub-domains that are used to assess an individual’s competency in tasks of personal and social sufficiency (Perry et al., 2009). The scores for each core domain are norm referenced and summed to generate an Adaptive Behavior composite (ABC) score that provides a holistic picture of an individual’s adaptive behavior across domains. The Vineland-3 tools were administered by three trained assessors at various timepoints in the participants’ lives, ranging from 1-23 years old. Individual assessor scores were cross validated with one another to ensure equivalency in assessment administration and scoring. Participant caregivers were those interviewed due to their proximity to and knowledge of the patients. Participants were given access to the Vineland assessment scores after they were generated.

### Analysis

The Principal Investigator and associated staff collected at least one Vineland-3 score for each participant over multiple years. Some participants were able to take the assessment more than once, thereby providing a longitudinal outlook. Natural history analysis and visualization was then performed using the GraphPad Prism software. All pathogenic variants were analyzed together and graphed by comparing their ABC standard score against the age at time of assessment. Further analysis was performed by filtering graphs by sex. All pathogenic variants, male participants, and female participants were also graphed according to their Communication (com), Daily Living Skills (dls), Social (soc), and Motor (mot) domain standard scores. The domain scores were further broken down into sub-domain standard scores to allow for more granular identification of participant strength and weakness over time. The Communication sub-domain scores were Receptive (rec), Expressive (exp), and Written (wrn). The Daily Living Skills sub-domains analyzed were Personal (per), Domestic (dom), and Community (cmm). The Socialization sub-domains of interest were Interpersonal Relationships (ipr), Play and Leisure (pla), and Coping Skills (cop). The Motor sub-domains of Interest were Fine Motor (fmo) and Gross Motor (gmo). Scores were only collected and included for this motor domain if patients were under the age of nine to stay within the validated age range of the exam (Perry et al., 2009), although it is possible to score older patients, just keeping this limitation in mind. Lastly, the Internalizing (int) and Externalizing (ext) sub-domain components of the Maladaptive Trait domain were also included in analysis, however the Critical Items component of the domain were not.

The scores were compared in Microsoft Excel with two-tailed equal variance t-tests to compare the adaptive behavior outcomes between males and females to determine if there were sex differences. The p-value was set at .05 for all calculations. Correlation coefficients were also calculated for each of the graphs made using Excel. A second round of calculations for the t-test and the correlation coefficients in the male cohort was performed to exclude NAA15-010 due to being three standard deviations above the mean in age compared to the other participants and having performed two standard deviations below the mean in Vineland ABC, Motor, Social, and Daily Living Skills standard scores. Data presented in tables includes calculations performed including NAA15-010 unless otherwise specified. The Vineland manual also provides Domain and ABC standard score ranges that correspond to adaptive behavior functioning level. “High functioning” individuals score between 130-140, “moderately high” between 115-129, “adequately” between 86-114, “moderately low” between 71-85, and “low functioning” less than 70. We clustered participants based off their scores and performed chi square statistics to determine if there was a further differentiation that could be made between the male and female cohort in how they function.

## Results

There were 27 participants who were administered the Vineland and included in this study. A breakdown of the different pathogenic variants in each sex can be seen in **Table 1**. In this cohort, there were 26 unique pathogenic variants identified. 25 of these variants were only present in one proband each. The p.His080Argfs*17 variant was present in two unrelated probands. Twenty-five participants developed their variant *de novo* and 2 participants have variants of unknown origin (due to the parents not being tested). Of the 27 participants, 9 are female and 18 are male with ages ranging from 1 year to 24.5 years (mean = 9.0 years, standard deviation (SD) = 5.2).

**Table 1.** NAA15 Pathogenic Variant Breakdown by Sex.

| Pathogenic Variant | Males | Females |
| --- | --- | --- |
| p.Arg27Gly | 1 |  |
| p.His80Argfs*17 | 2 |  |
| p.Trp83X |  | 1 |
| p.Asn113Ser |  | 1 |
| p.Glu173* | 1 |  |
| p.Glu173NfsX54 | 1 |  |
| p.Tyr333* | 1 |  |
| p.Asp335Glufs*7 | 1 |  |
| p.Asn362MetfsX36 |  | 1 |
| p.Cys484Arg | 1 |  |
| p.Arg549X | 1 |  |
| p.Ala585Thr | 1 |  |
| p.Gln621* | 1 |  |
| p.Asn623Ilefs* | 1 |  |
| p.Lys628Glu | 1 |  |
| p.Ala678Cysfs*3 | 1 |  |
| p.Tyr682* |  | 1 |
| p.Ala719Thr | 1 |  |
| p.Arg782X |  | 1 |
| p.Ile796Thr | 1 |  |
| p.Asn864Ser |  | 1 |
| c.0692-1G>T (intronic) | 1 |  |
| c.139+2T>A: IVS2+2T>A (intronic) |  | 1 |
| c.1087+2T>C (intronic) |  | 1 |
| c.1753+1G>A (splice site) | 1 |  |
| c.2057-2A>G (splice site) |  | 1 |
| **Total: 27** | **18** | **9** |

Compared to the general population, individuals with *NAA15* variants scored below average on the Vineland-3 Assessment (where average is mean=100, sd=15). This was true for ABC standard scores as well as the main domain scores. The average ABC score among *NAA15* variants was 68.5 (SD = 22.1). Females tended to outperform males across adaptive behavior domains. However, except for the Motor domain score, these differences were not statistically significant. A summary of the Vineland 3 scores can be seen in **Table 2**.

**Table 2.** Vineland ABC and Domain Standard Scores by Sex.

|  | Communication | Daily Living Skills | Social | Motor | ABC Standard |
| --- | --- | --- | --- | --- | --- |
| **Total Average** | **64.1** | **70.2** | **72.7** | **71.1** | **68.5** |
| SD | 26.7 | 21.1 | 23.9 | 17.5 | 22.1 |
| **Female Average** | **70.0** | **75.4** | **78.2** | **83.2** | **74.9** |
| SD | 28.4 | 22.9 | 23.8 | 11.6 | 24.4 |
| **Male Average** | **60.9** | **67.4** | **69.6** | **63.9** | **65.0** |
| SD | 25.1 | 19.5 | 23.3 | 16.4 | 20.0 |
| **t test** | **0.27** | **0.22** | **0.24** | **0.01\*** | **0.15** |

\*Asterisk denotes significance (p<.05)

Graphical representations of the ABC standard scores over time can be seen in **Figure 1**. Before stratifying by sex, there appears to be an overall downward trend in score with age. **Figure 2** shows the ABC standard scores separated by male and female. Separation by sex shows a moderate linear increase in adaptive function in males over time, when excluding NAA15-010, whereas females appear to show a moderate linear decay. There is also a moderate linear decay in the female Daily Living Skills domain over times whereas the males showed a moderate positive linear growth when excluding NAA15-010. **Table 3** showcases the remaining calculated correlation coefficient values for the Vineland ABC and Domain standard scores for the females, males, and males excluding NAA15-010.

**Figure 1.**
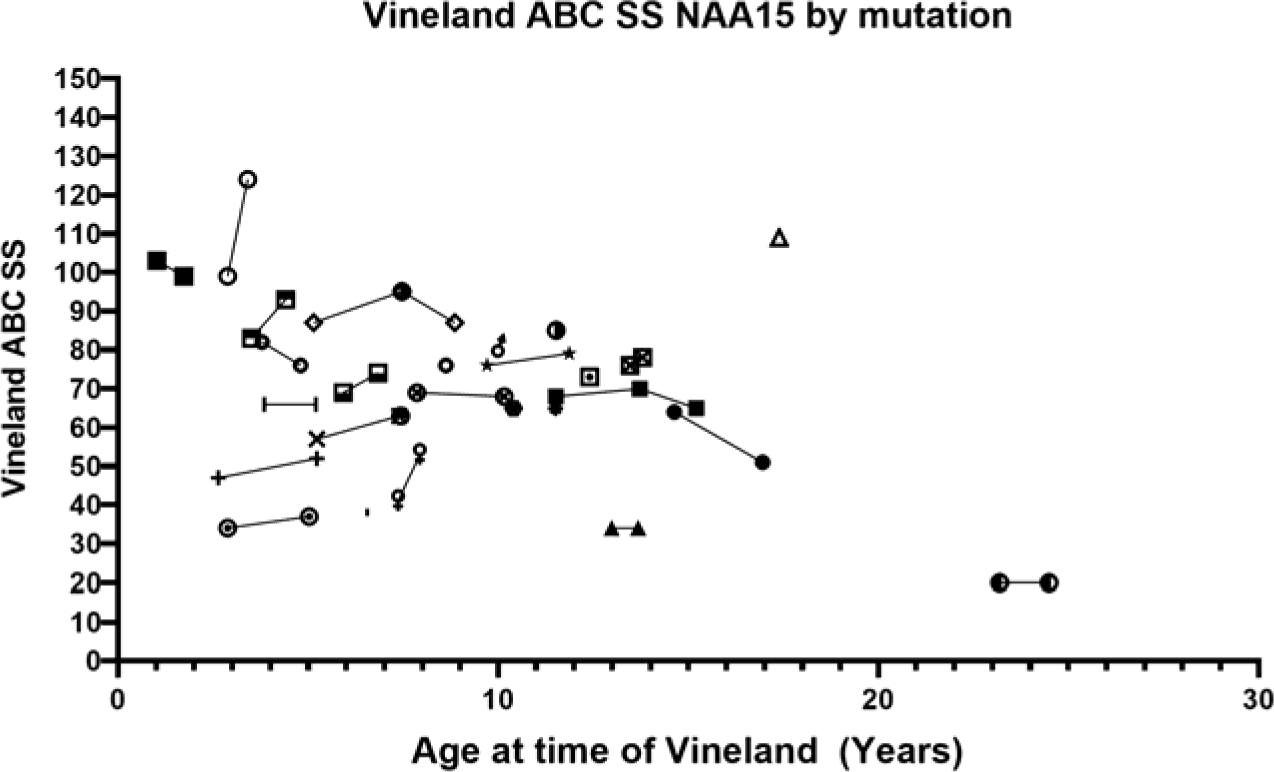
ABC Standard Score vs Age at time of Vineland Administration. Each point on the graph represents a single participants ABC standard score at a given time point. Points connected by lines show change in single participant scores over time.

**Figure 2.**
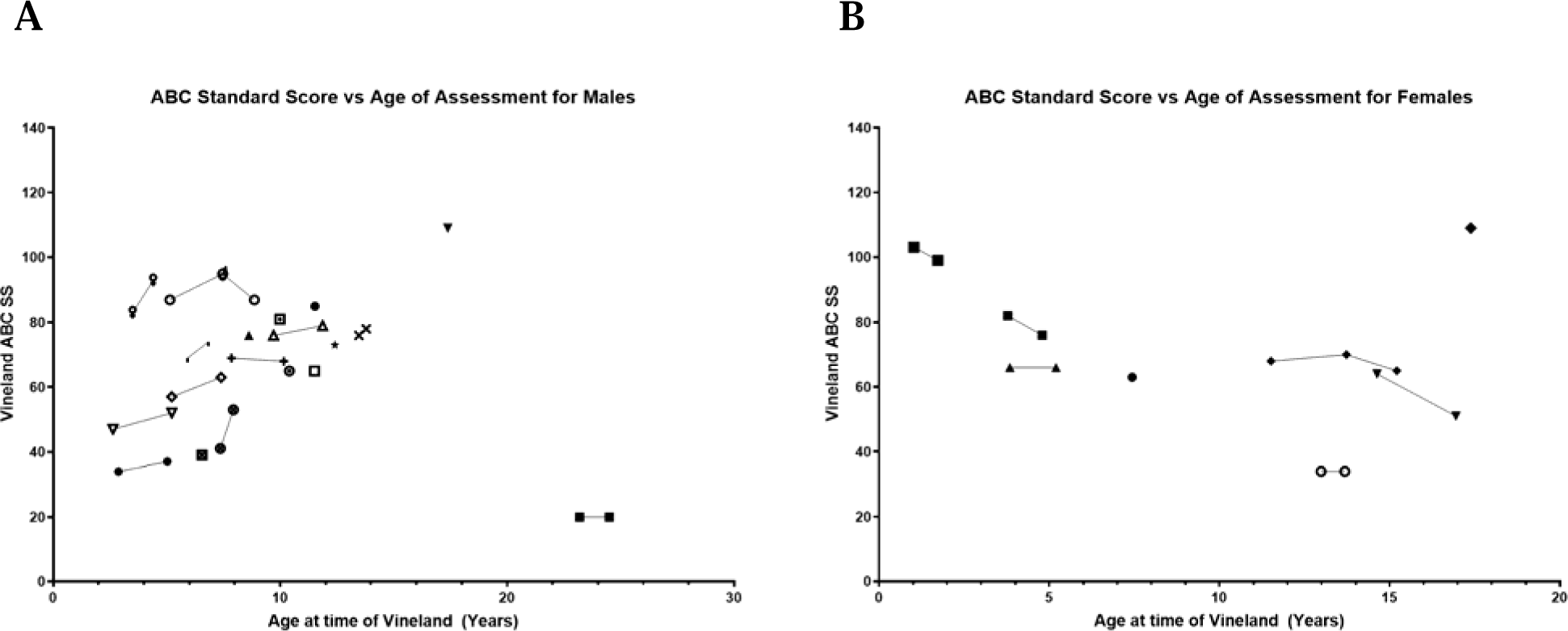
ABC Standard Score vs Age at time of Vineland Administration by Sex. Each point on the graph represents a single participants ABC standard score at a given time point. Points connected by lines show change in single participant scores over time.

**Table 3.** Correlation Coefficients of Vineland Domain Standard Scores vs Age.

|  | Communication | Daily Living Skills | Social | Motor | ABC |
| --- | --- | --- | --- | --- | --- |
| Female Correlation Coefficient | -0.49 | -0.58 | -0.46 | -0.71 | -0.51 |
| Male Correlation Coefficient | -0.15 | -0.27 | -0.31 | 0.15 | -0.28 |
| Male Correlation Coefficient excluding ID: NAA15-010 | 0.35 | 0.49 | 0.26 | 0.15 | 0.38 |

The Communication, Daily Living Skills, Social, and Motor standard score graphs can be seen in **Figure 3** and follow similar trends for females and males, when excluding NAA15-010, to the ABC scores over time. It should be noted that excluding NAA15-010 did not significantly change the t-test statistics of the standard scores when comparing the males and the females. The magnitude at which the females outperformed the males decreased. However, except for the Motor domain score, their differences were still not significant.

**Figure 3.**
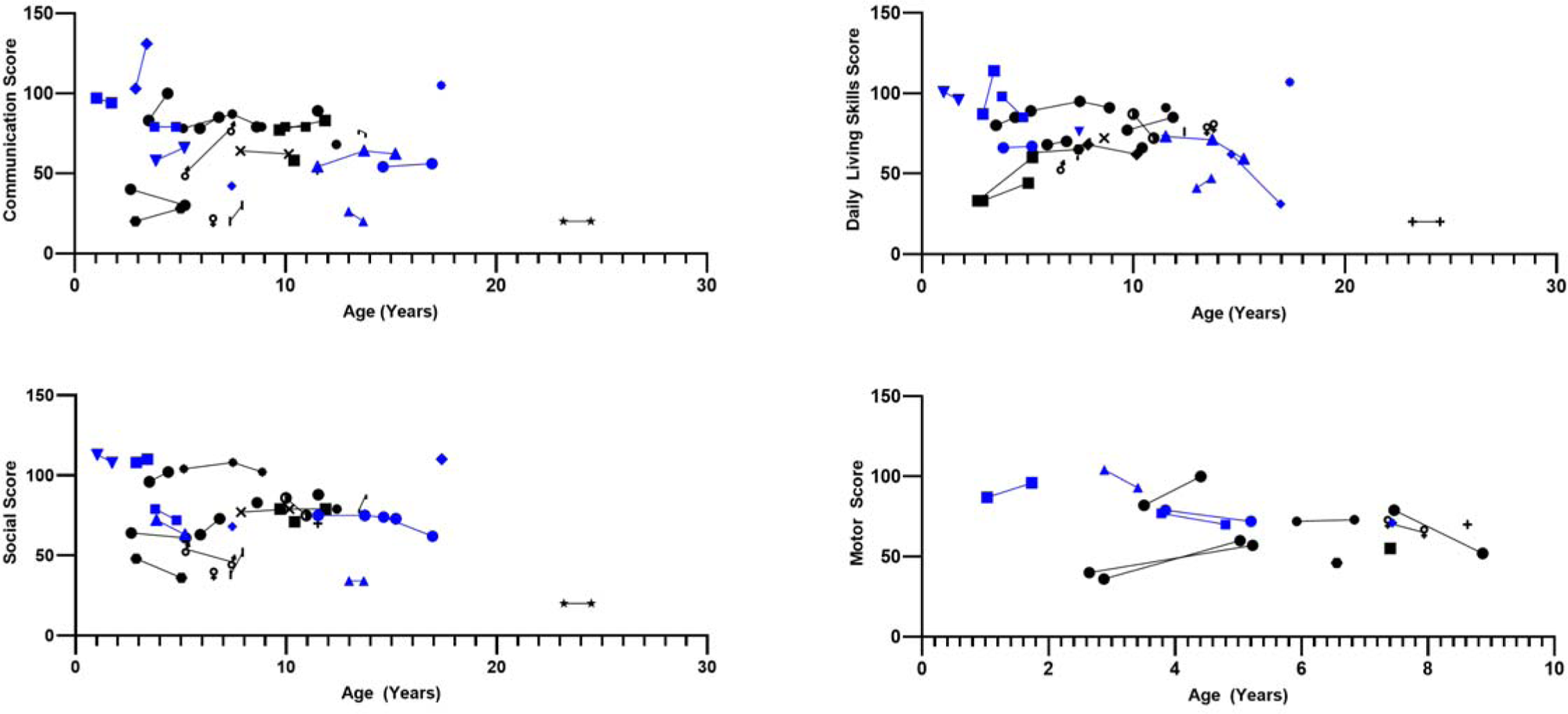
Vineland Domain Score vs Age at time of Vineland Assessment. Points in black represent Males. Points in Pink represent Females. Each point on the graph represents a single participants Vineland Domain standard score at a given time point. Points connected by lines show change in single participant scores over time.

Dividing the Motor domain into its Fine Motor and Gross Motor sub-domains elucidated the difference between sexes. Males, with and without the inclusion of NAA15-010, scored 7.8 (SD = 3.2) on average on the Fine Motor subdomain component of the Vineland. This is significantly less than the females, who had an average score of 13.3 (SD = 3.5, p = .001). On average for Gross Motor subdomain scores, males (mean = 8.5, SD = 4.0) also performed worse than their female counterparts (mean = 10.4, SD = 2.2), although this difference was not statistically significant. There were no other significant differences between how the sexes performed in the various sub-domains. A summary of the breakdown of each domain into its sub-domain components can be seen in **Table 4**.

**Table 4.**
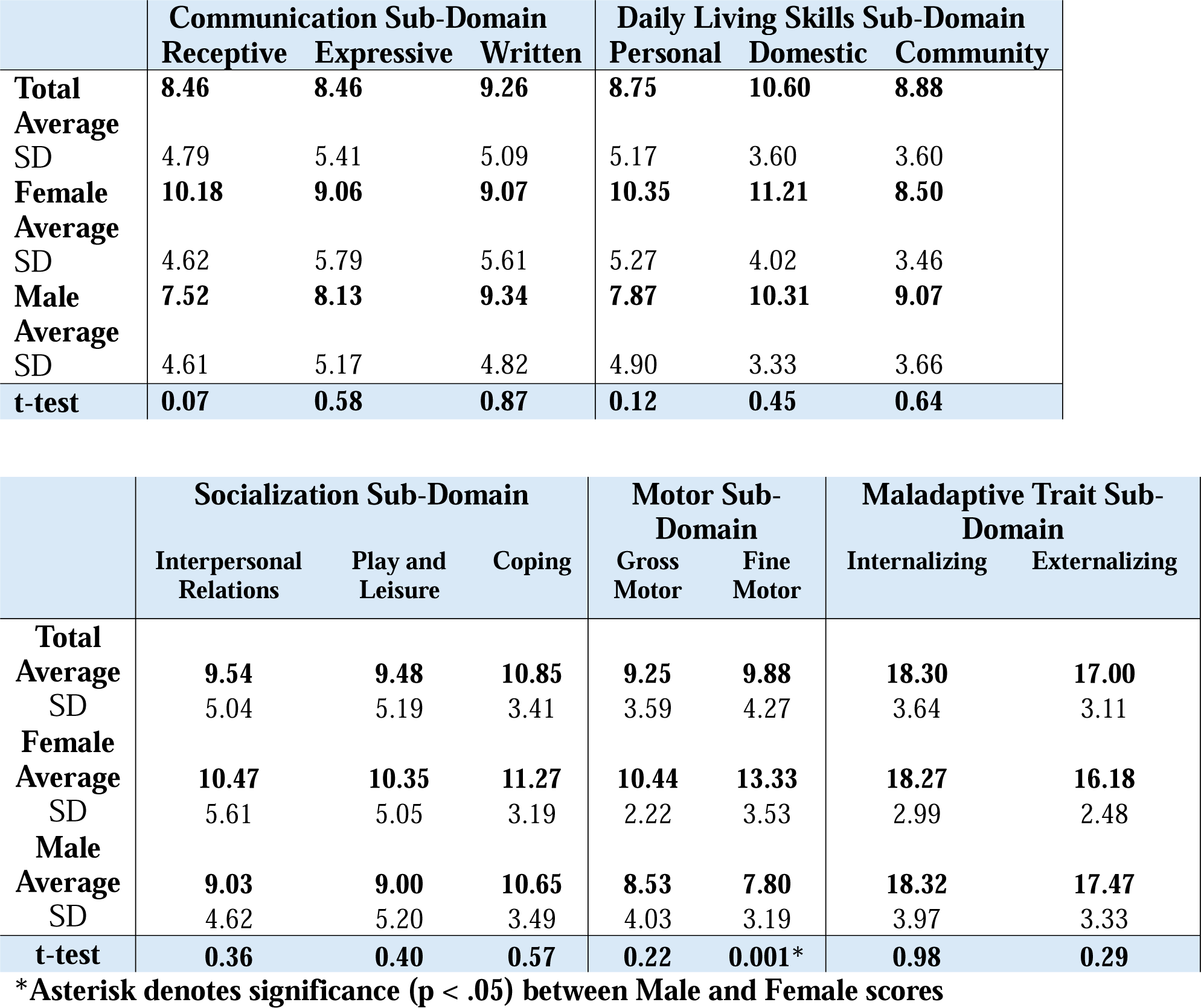
Vineland Sub-Domain Standard Scores by Domain and Sex.

Each domain sub-domain score can be seen in **Figure 4**. The strongest negative linear associations exist between the Coping, Fine Motor, and Domestic sub-domains over time in the females with *NAA15* variants. There are also moderately strong negative linear associations in the Receptive and Personal sub domains. The correlation coefficients in the males were not as strong as those in the females. However, when excluding NAA15-010, the Personal sub-domain had a moderate positive linear relationship over time. Calculated correlation coefficients for each sub-domain can be seen in **Table 5**.

**Figure 4.**
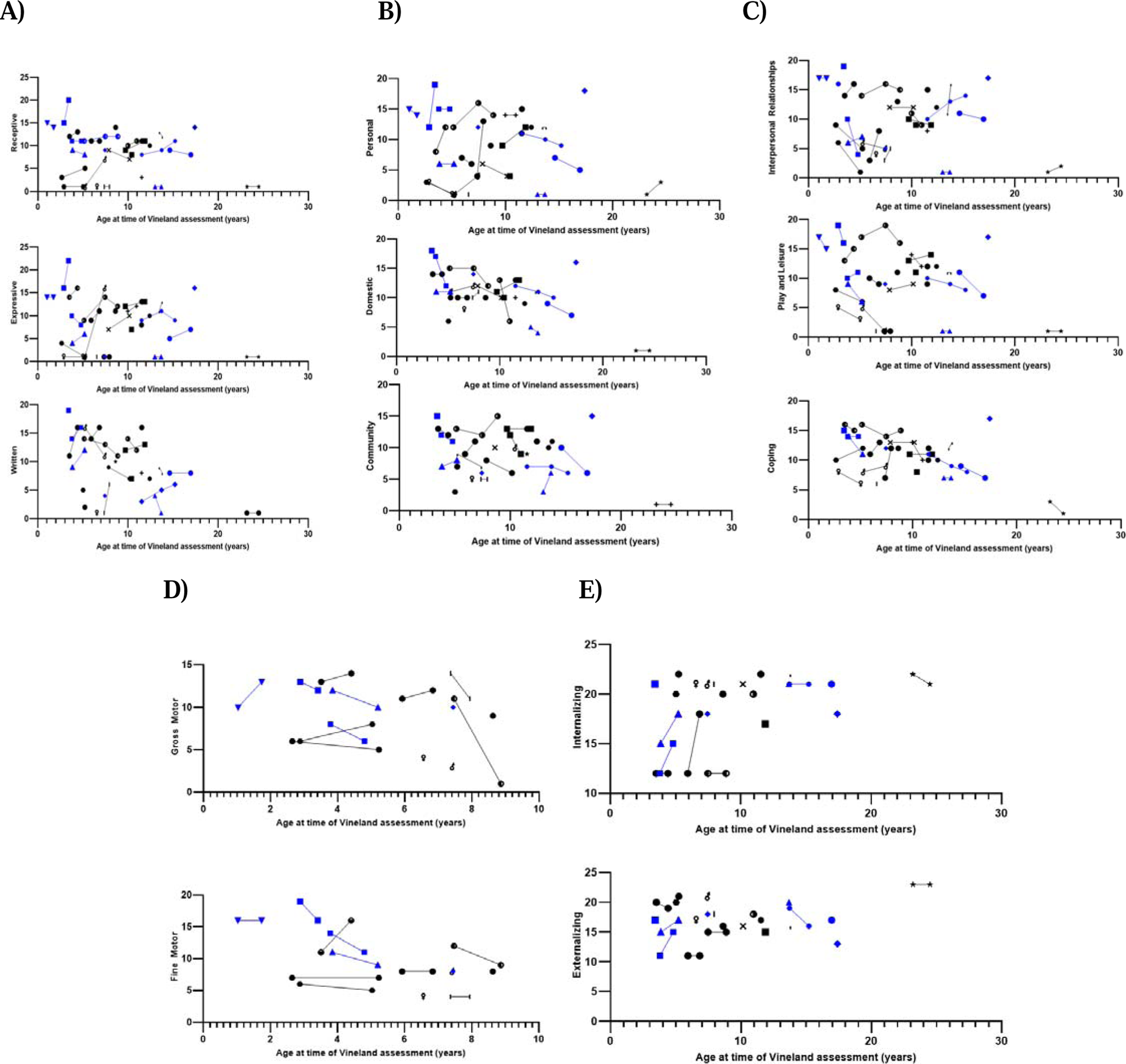
Vineland Sub-Domain Score vs Age at time of Assessment. A) Communication Sub-Domain Standard Scores. B) Daily Living Skills Sub-Domain Standard Scores. C) Socialization Sub-Domain Standard Scores. D) Motor Sub-Domain Standard Scores E) Maladaptive Trait Standard Scores. Points in black represent Males. Points in Blue represent Females. Each point on the graph represents a single participants sub-domain standard score at a given time point. Points connected by lines show change in single participant scores over time.

**Table 5.**
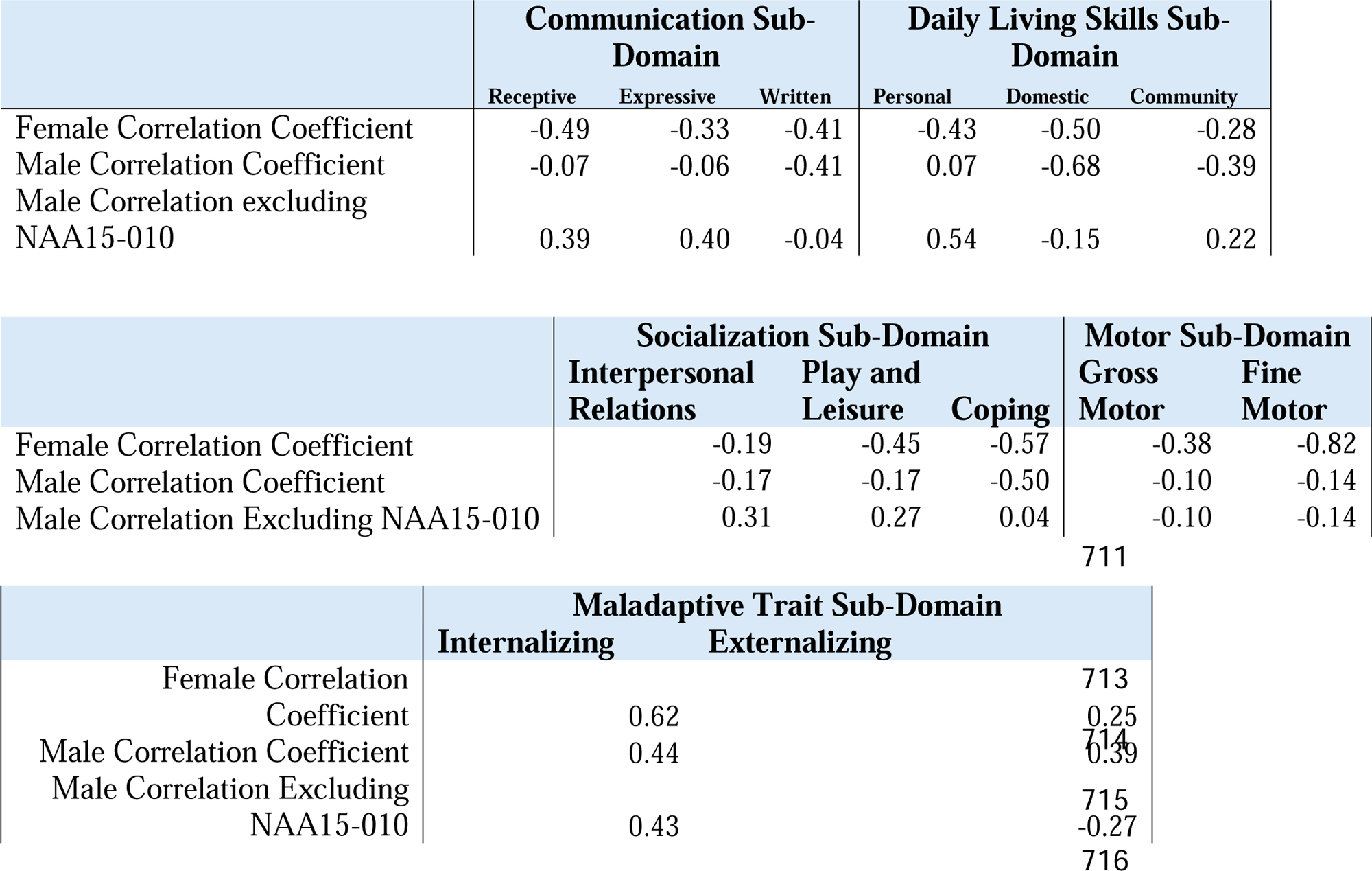
Correlation Coefficients of Vineland Sub-Domain Standard Scores vs Age.

Categorical analysis based on the Vineland manual category designations can be found in **Table 6**, comparing males versus females. There was a significant difference between males and females for performance on the Motor domain (p = .02). The remaining comparisons were not significant.

**Table 6.** Comparison of Vineland Adaptive Behavior Categories by Sex per Domain.

|  | Communication |  | Daily Living Skills |  | Socialization |  | Motor |  | ABC |  |
| --- | --- | --- | --- | --- | --- | --- | --- | --- | --- | --- |
|  | Male | Female | Male | Female | Male | Female | Male | Female | Male | Female |
| Moderately High | 0 | 0 | 0 | 0 | 0 | 0 | 0 | 0 | 0 | 1 |
| Adequate | 3 | 4 | 5 | 6 | 8 | 5 | 1 | 4 | 4 | 4 |
| Moderately Low | 13 | 2 | 11 | 4 | 10 | 7 | 5 | 4 | 11 | 2 |
| Low | 12 | 10 | 13 | 7 | 10 | 5 | 9 | 1 | 13 | 10 |
| $\chi^2$ | 0.06 | | 0.42 | | 0.92 | | 0.02* | | 0.11 | |

\*Asterisk denotes significance (p < .05) between Male and Female scores

According to NAA15-010’s caregiver, they were diagnosed with focal seizures in early adolescence **(Supplemental 1)**. In conjunction to the focal seizure diagnosis, the patient developed catatonia after chronic mold exposure leading to a leading diagnosis of Pediatric Acute-onset Neuropsychiatric Syndrome (PANS) (Gagliano et al., 2023). The caregivers ordered independent mold testing of their residence **(Supplemental 2)** and a urine fungal toxin panel **(Supplemental 3)** in hopes of providing additional evidence to support the diagnosis. According to the fungal toxin panel, the levels of gliotoxin and trichothecene were present in the 50^th^-99^th^ percentile. While the lab did not reference the range at which the percentiles were set, gliotoxin is associated with *Aspergillus* and trichothecene with *Fusarium* (Janik et al., 2021; Ye et al., 2021). The lack of treatment received for their PANS-associated catatonia over an approximately ten-year period is NAA15-010’s caregiver’s explanation for their stark decrease in function **(Supplemental 1**).

NAA15-034, in contrast to NAA15-010, performed better on the Vineland-3 assessments compared to the rest of the tested cohort. NAA15-034 did not have childhood exposure to mold or other toxins, according to caregiver reports, but does have a history of hyperbaric oxygen therapy (HBOT) (two sessions daily) that was started on the recommendation of clinicians who believed the proband had intellectual disability with static encephalopathy or possibly a mitochondrial disease. NAA15-034 had her first session at before the age of 2 years, at which time she was, according to correspondence with the caregivers, one year behind developmentally, being unable to crawl or support weight on her legs. On day 4, after the 8th session, NAA15-034 crawled and was able to bear weight on her legs for the first time. NAA15-034 continued to have HBOT sessions until around 5 years old. At that time, she had caught up developmentally with their peer group. Further information regarding NAA15-034, and their disease progression and associated symptoms, is currently being written up in a case report highlighting their progress with HBOT.

## Discussion

Individuals with *NAA15*-related neurodevelopmental syndrome performed significantly worse than the mean on the Vineland-3. This difference was expected, as the expression of the NatA complex is ubiquitous across tissue types and plays a prominent role in development (Cheng et al., 2020; Liszczak et al., 2013; Lyon et al., 2023). However, individuals with *NAA15* variants (n=27, mean=68.5) perform significantly better, on average, than individuals with *NAA10* variants (n=58, ABC mean=40.4) (Makwana et al., 2024). NAA10 is the catalytic subunit of the NatA complex where NAA15 is an auxiliary subunit. In the presence of NAA15, NAA10 localizes to the ribosome and preferentially acetylates N-termini Ser, Ala, Thr, Val, and Gly (Arnesen et al., 2005; Park & Szostak, 1992). In the absence of NAA15, NAA10 has been shown to acetylate acidic N-termini residues and localize to the cytoplasm suggesting additional function independent of the NatA complex (Liszczak et al., 2013; Van Damme et al., 2011). NAA10 has also been shown *in vitro* to affect dendritic arborization, such that both over and under expression of the protein leads to abnormal dendritic development (Chou et al., 2024; Ohkawa et al., 2008). Animal models have shown that abnormalities in dendritic arborization during development can lead to Autism Spectrum Disorder-like behavior suggesting NAA10 specifically may play an even larger role in cognitive development by itself than as part of the NatA complex (Barón-Mendoza et al., 2021; Martínez-Cerdeño, 2017). Given the greater functional significance of a working NAA10 protein, it stands to reason that *NAA10* related neurodevelopmental syndrome would have a more severe presentation than that of *NAA15* related neurodevelopmental syndrome.

When comparing performance by sex, the females with *NAA15* variants, on average, performed better than the males. Except for two participants with an unknown inheritance pattern, all *NAA15* pathogenic variants in this cohort occurred *de novo*. The difference in sex is unexpected, however, it could be due to small sample sizes. Additionally, given that 23 out of 25 pathogenic variants in this cohort are unique (**Table 1**), it was not possible to assess whether the differences between sexes are significant at a genotype level. Further research with a larger cohort should be conducted to identify recurrent pathogenic variants and their impacts, if any, on functional and cognitive outcomes.

While females performed, on average, better than males across all adaptive domains, they performed significantly better on the motor domain and fine motor sub-domain portions of the Vineland. Pathogenic variants in *NAA15* have been associated with infant onset dystonia in a few individuals (Cheng et al., 2018; Danti et al., 2024; Lyon et al., 2023; Yubero et al., 2022), which could be one reason why the cohort performed worse on motor tasks compared to the norm population. However, there did not appear to be a difference in the presentation of musculoskeletal manifestations between the sexes in the referenced literature. Furthermore, there is a case where an *NAA15* variant led to adult-onset parkinsonism in a man presenting with other manifestations of *NAA15*-related neurodevelopmental syndrome, such as speech delay and intellectual disability (Straka et al., 2022). Continued genetic testing of individuals with adult-onset dystonia and longitudinal monitoring of the current cohort for motor disorders would be necessary to help determine if the difference in motor function between male and females is due to chance, sex differences, or pathogenic variant type.

In aggregate, it is difficult to determine if there is an overall increase or decrease in function over time in the participants. However, when comparing performance by sex, there is a negative correlation between adaptive behavior in females and age. The decline with age has the strongest correlation between the coping, domestic, and fine motor sub-domains. With a decline in fine motor capacity, it would stand to reason that an individual’s ability to take care of oneself (domestic) would also decline. However, both the domestic and fine motor sub-domains have age restrictions leading to unequal administration of those portions of the exam creating an even smaller population size to compare from (Farmer et al., 2020; Perry et al., 2009, p. 3). Furthermore, this decline in function contradicts the mild recovery in gross motor, personal-social, and language development function previously reported in the literature (Cheng et al., 2018, 2020; Tian et al., 2022). However, some previous reports of mild functional recovery, such as that by Tian et al. in 2022, were limited by small sample size (n=4). This study also used the Children Neuropsychological and Behavior Scale-Revision (CNBS-R2016) and Ages and Stages Questionnaires (ASQ), rather than Vineland-3, making direct comparisons more difficult. The decline in the coping sub-domain in the females could also be partially explained by the decrease in motor skills, as there is evidence that similar motor declines in those with Autism Spectrum Disorder (ASD) have led to decreased coping and socialization skills overall (Peyre et al., 2024; Raditha et al., 2023). Additionally, several patients in this cohort ranged from ages 11 to 17, a time period that is traditionally tumultuous for adolescents due to various physical and hormonal changes, and is associated with increased stress for those with ASD (Cridland et al., 2014; Steward et al., 2018; Tager-Flusberg & Kasari, 2013). Ultimately, follow up studies need to be performed on this cohort to determine if they are continuing to show decline over time, if there is a bottoming-out effect, or if they begin to improve after navigating adolescence.

In contrast to the female decline over time, males when analyzed without NAA15-010 have a moderate increase in adaptive function with age. However, there are not strong correlations present in any subsequent sub-domain analysis performed. Additionally, when including NAA15-010, there were no or small correlations between Vineland score and age.

NAA15-010 was removed from the analysis due to their having performed much worse than the rest of their cohort. One explanation for their decreased performance could be due to their focal seizures as there is evidence suggesting adolescent onset seizures are associated with nonspecific neurodevelopmental declines (Dreier et al., 2019; Nickels et al., 2016). Another explanation for the decrease in performance could be the presence of *Aspergillus* and *Fusarium* in the proband’s urine. Various case reports have suggested that there is a link between the presence of *Aspergillus* and Autism Spectrum Disorder (Baker & Shaw, 2020; Markova, 2019) and schizophrenia (Bettoni et al., 1984). Thus, while there are no reported cases of PANS associated with *Aspergillus* colonization, the diagnosis could explain both the proband’s catatonia and decline (Rogers et al., 2019). This once again highlights that the trajectory of any genetic disease, including “Mendelian” ones, can be dramatically altered by environmental, other genetic, or stochastic influences (Lyon & O’Rawe, 2015, pp. 289–318).

Despite NAA15-010 and NAA15-034 being two of the oldest probands in the cohort, NAA15-034 markedly outperformed NAA15-010 across Vineland domains. They also performed markedly better than the rest of the study cohort. An explanation for this could be the multiple HBOT sessions NAA15-034 underwent throughout development. HBOT is clinically indicated for various conditions such as gas emboli, thermal burn injury, carbon monoxide poisoning, and central retinal artery occlusion among others (Ortega et al., 2021). The therapy acts to increase angiogenesis and wound healing, exerts antimicrobial effects, and helps rapidly alter circulating O2 levels (Ortega et al., 2021). HBOT also has some theoretical indications for use in cancer treatment (Lu et al., 2019; Thews & Vaupel, 2015), immune modulation (Novak et al., 2016; Resanovic et al., 2019), and as a neuroprotective agent (Chazalviel et al., 2016; Zhou et al., 2016). A clinical trial has shown that HBOT may have some efficacy in improving symptoms in pediatric post-concussive syndrome (Hadanny et al., 2022). Multiple trials, however, have shown little evidence for HBOT in improving adaptive behavior in autism spectrum disorder (ASD) (Bent et al., 2012; El-Tellawy et al., 2022; Sampanthavivat et al., 2012). NAA15-related neurodevelopmental disorder presents with similar behavioral deficits to ASD, but has a known genetic deficit whereas ASD represents a spectrum of different etiologies leading to a similar over-arching cognitive phenotype. This difference may explain the lack of HBOT effect in traditional ASD cohorts. Future work could consider treating any NAA15 mouse models with HBOT to better understand the mechanism of effect and to identify an initial safety profile.

The overall decrease in female adaptive behavior scores and the overall significantly decreased male adaptive scores suggest that there is an urgent need for additional studies and development of interventions for *NAA15*-related neurodevelopmental disorder. Future research should aim to complete the clinical timeline as best as possible. The present cohort only included individuals up to the age of 25 due to the rarity of the disease. Without insights into the later stages of the disorder, it is more difficult for clinicians to guide caregivers on what to expect at different stages of a patient’s life.

*NAA15*-related neurodevelopmental disorder is thought to be caused by haploinsufficiency (Cheng et al., 2018; Tian et al., 2022). For this mechanism of disease, there are several therapies in development that show promise in cell lines and animal models in similar disorders, such as Rett Syndrome and Angelman Syndrome, to help restore protein function and dosage to adequate levels (Albadri et al., 2017; Burbano et al., 2022; Hill & Meisler, 2021; Milazzo et al., 2021; Palmieri et al., 2023; Protic et al., 2019). More recently, a case of hereditary spastic paraplegia type 50 was treated with intrathecal delivery of an AAV9 encoded viral vector. Investment in similar interventions for NAA15-related neurodevelopmental syndrome, if implemented early enough, may restore protein function and restore adaptive behavior functioning in affected patients (Dowling et al., 2024).

## Conclusion

*NAA15*-related neurodevelopmental disorder is closely related to Ogden Syndrome (*NAA10*-related neurodevelopmental disorder), as both the NAA15 and NAA10 proteins are part of the NatA complex. NAA15 plays a supporting role by helping to bring the complex to the ribosome, with dysfunction leading to clinical manifestations in the heart, musculoskeletal system, and brain. The pathogenic variants in *NAA15*-related neurodevelopmental disorder are heterozygous, so there is still one functioning copy of *NAA15*, meaning that the NatA complex can still form, but likely with haploinsufficiency. As such, *NAA15* pathogenic variants usually present less severely than those with pathogenic *NAA10* variants. Repeated Vineland-3 administration to a cohort of 27 patients with unique *de novo NAA15* pathogenic variants has revealed the natural history of the disease, showcasing an overall decrease in adaptive functioning compared to the norm from infancy till adolescence. Females tended to perform on average better than males. However, over time, females showed a decline in function whereas males showed an increase. Additional natural history studies need to be performed to understand disease outcomes outside of adolescence, difference between sexes, and to identify outliers who perform significantly better or worse than their peers, so genetic or environmental factors influencing disease can be identified.

## Supplemental Information

**S1. NAA15-010 Caregiver Report**

**S2. Anonymized Mold Report**

**S3. Anonymized Fungal Toxin Report**

## Author Contributions

GJL and RH conducted all virtual interviews with participants and were responsible for primary Vineland data collection, with data curation conducted by EM. RM and CC were responsible for data analysis and project conception, along with GJL. The first draft of the manuscript was written by RM and CC, with critical revision performed by GJL and RP at several points.

## Acknowledgments

We thank the families and the foundation, Ogden CARES for their participation and support. We thank Ellen Israel for assistance with data collection early on in the project.

## Ethical Approval

Both oral and written patient consent were obtained for research and publication, with approval of protocol #7659 for the Jervis Clinic by the New York State Psychiatric Institute Institutional Review Board.

## Data Availability

All data are deidentified to protect patient privacy, and the underlying data cannot be shared due to these same privacy restrictions.

